# Validation of the Behavioural and Social Drivers of Influenza Vaccination Survey in Australia: Cognitive Interviews and Psychometric Evaluation

**DOI:** 10.64898/2025.12.08.25341812

**Authors:** Majdi M. Sabahelzain, Maria Christou-Ergos, Julie Leask

## Abstract

**Background:** Unvaccinated individuals are at greater risk of seasonal influenza. In Australia, vaccination coverage is consistently low among the most at risk groups eligible for free influenza vaccine. To establish an annual national monitoring of the behavioural and social drivers of influenza vaccination, we validated the World Health Organization’s Behavioural and Social Drivers of Influenza Vaccination survey. This is the first study reporting the validation results of the newly developed survey.

**Methods:** Two stages of validation occurred. First, we conducted cognitive interviews and revised survey items based on feedback. Then, we collected quantitative data and performed psychometric analysis, including explanatory and confirmatory factor analysis. We also assessed the association between the survey items and the intention to receive an influenza vaccine using polychoric correlation and used a random forest model to identify the importance of each item in predicting the intention to receive an influenza vaccine.

**Results:** We modified nine of the 31 survey items based on cognitive interviews with 16 adults. Based on the psychometric evaluation of 2055 respondents, we identified and validated five factors that underlie the survey structure and aligned well with the domains of the BeSD framework: Thinking and Feeling, Social Processes, Motivation, and Practical Issues. Each item correlated with the intention to receive influenza vaccine, particularly those related to perceived vaccine importance and social responsibility. Only one item - autonomy of decision-making about influenza vaccination - showed no significant association with the intention to receive influenza vaccination and no significant correlation with its underlying factor.

**Conclusion:** The cognitive interviews and psychometric evaluation of the survey showed strong validity in an Australian population. We recommend further validation of the survey in various contexts, with a particular focus on assessing its predictive validity with actual vaccination uptake.

## 1. Background

Influenza presents a significant public health challenge, leading to considerable morbidity and mortality annually [1]. It is estimated that unvaccinated individuals have a higher risk of seasonal influenza infection, with nearly 1 in 5 unvaccinated children and 1 in 10 unvaccinated adults estimated to contract the infection each year [2]. In Australia, the disease is characterised by seasonal patterns and the potential for severe complications, especially among vulnerable populations such as adults aged 65 and over, children aged 5 and under, pregnant people, individuals with chronic health conditions, as well as Aboriginal and Torres Strait Islanders [3]. Although the influenza vaccine remains the most effective intervention to reduce the morbidity and mortality of the disease [1] and is being provided free of charge to these groups in Australia, vaccination coverage is consistently low among these target populations [4].

Understanding the underlying behavioural and social drivers of influenza vaccine uptake is essential for developing effective interventions to increase vaccination coverage [5,6]. The World Health Organization (WHO) recommends all countries regularly collect quantitative and qualitative data using Behavioural and Social Drivers of Vaccination (BeSD) surveys and priority indicators, focusing on areas and sub-populations that have gaps in vaccination coverage and inequities [7]. The BeSD survey is a globally standardized and validated survey designed to measure modifiable beliefs and experiences related to vaccination measured within four domains, including thinking and feeling about vaccination, social processes, motivation, and practical issues. Some studies have used individuals’ intentions, willingness, and hesitancy to receive vaccinations as outcome indicators [8], particularly when it is not feasible to collect data on current vaccination status. These indicators are part of the motivation domain in the BeSD [7].

In this context, WHO has also developed the Behavioural and Social Drivers of Influenza Vaccination tools [9]. These tools were originally based on the BeSD tools of childhood and COVID-19 vaccination [7]. The survey was developed by an expert group and informed by a literature review and prior studies of BeSD tools in different contexts [10]. Questions have now been adapted to assess influenza vaccination.

The current study is part of a larger research project using mixed methods aiming to provide a foundation for annual national monitoring of the behavioural and social drivers of influenza vaccination. We followed the World Health Organization’s recommendation to assess the new and adapted tools [9]. To establish content validity and determine item clarity, we conducted cognitive interviews to refine the survey questions for the Australian context. A modified survey was delivered in the field. From the responses, we performed a psychometric analysis to evaluate the survey items in Australia. This is the first study reporting the validation results on the newly developed survey.

## 2. Methods

This paper describes two stages of validation: cognitive interviews and psychometric evaluation of the survey items. First, we conducted cognitive interviews and revised the survey items based on the feedback. Subsequently, we collected quantitative data using the revised items and performed psychometric analysis.

### 2.1. Cognitive interviews

Cognitive interviews were conducted to pre-test survey questions and ensure that items generated the intended information and that response options were appropriate. This allowed researchers to adjust and refine the draft questions ahead of psychometric evaluation. Cognitive interviews sought to address issues with:

- Language – how people interpret and understand words or phrases.
- Inclusion –whether certain concepts are considered within the intended scope of the question.
- Temporal patterns – the time period that applies to a question.
- Computation – mental arithmetic required to answer a question.

#### 2.1.1. Sample and recruitment

Adults from across Australia were recruited through an external agency. An online advertisement was distributed to their panel, and those who agreed to participate provided written consent and arranged a time for an online interview with a researcher.

#### 2.1.2. Data collection and measures

During the interview, the researcher read aloud each survey question which was also presented visually to the participant via a shared screen. The researcher recorded the interviewees’ responses to the survey question before asking them between one and three questions designed to ascertain how they understood the meaning of the question and how they interpreted and answered it. Some examples of common cognitive interview questions used were: “How did you decide on your answer to this question?” and “Did the response options fit with the answers you wanted to give?”. For some questions, a visual analogue scale was employed to help quantify the participant’s responses, as shown in Figure 1.

**Figure 1:**
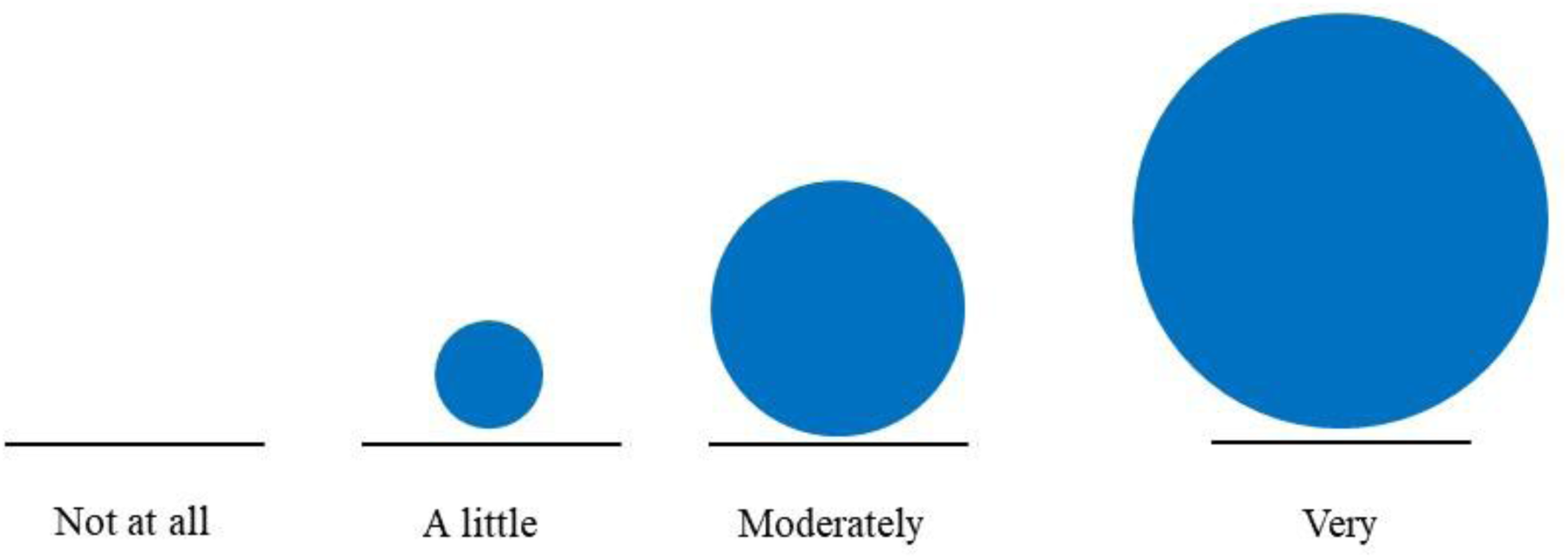
Visual analogue scale presented to participants [7].

#### 2.1.3. Data analysis

The researcher took extensive notes during the cognitive interviews. Following the completion of each interview, the interviewer recorded field notes regarding how the interview progressed and recorded any issues or observations related to the performance of the survey items. Responses and notes relating to each question were combined into a spreadsheet organised by item. Members of the research team engaged in regular meetings to discuss findings as they emerged. Careful inspection of each item and its associated notes revealed issues that were either (i) repeatedly observed across many participants or (ii) affected few participants but still threatened validity. Once cognitive interviews were completed, researchers collectively decided how to improve the validity of survey items for use in the Australian context.

### 2.2. Psychometric evaluation of survey items

#### 2.2.1. Sample and recruitment

Participants were recruited from an external panel provider to create a sample of 2,055 Australian adults over 18 years of age. The sample aimed to reflect proportions in the national population based on age, gender, and location.

#### 2.2.2. Data collection and measures

Data were collected using an online survey in March 2024. The WHO’s Behavioural and Social Drivers of Vaccination survey tool, adapted for influenza vaccination, was administered. Annex 1 lists the full set of items in the survey.

#### 2.2.3. Data analysis

SPSS version 29 was used to generate frequencies of survey items and perform explanatory factor analysis (EFA) to explore the structure of the newly developed survey. Confirmatory factor analysis (CFA) was also conducted with an oblique rotation (Promax) on the set of survey items using SPSS Amos v29. The names of the underlying factors were refined and agreed upon through a structured discussion and consensus between the co-authors, ensuring their consistency with the theoretical construct of BeSD. To assess criterion validity, the bivariate correlation was conducted using the Polychoric correlation command on Stata SE18. This assessed the association between the survey items and the intention to receive the influenza vaccine.

A random forest model was computed using the ’forest’ package and command in Stata SE18 to determine the importance of each item in predicting the intention to receive the influenza vaccine within the full set of items. Random Forest is a machine-learning algorithm for identifying which predictors were highly predictive of a dependent variable. This analysis is different from bivariate analyses. The syntax for the ’rforest’ command was obtained from Schonlau et al. [11].

From a total of 31 survey items, 21 were included in the psychometric analysis (Table 2). The item for vaccination uptake/status (VS1), items with multiple options (SP6, P02, P04, P08, P10) and those not applicable to all participants (VS2, VS3, VS3b, and P09) were also excluded.

## 3. Results

### 3.1. Cognitive interviews

A total of 16 adults from across Australia were recruited to participate in an interview. Table 1 summarises the characteristics of participants in the cognitive interviews.

**Table 1:**
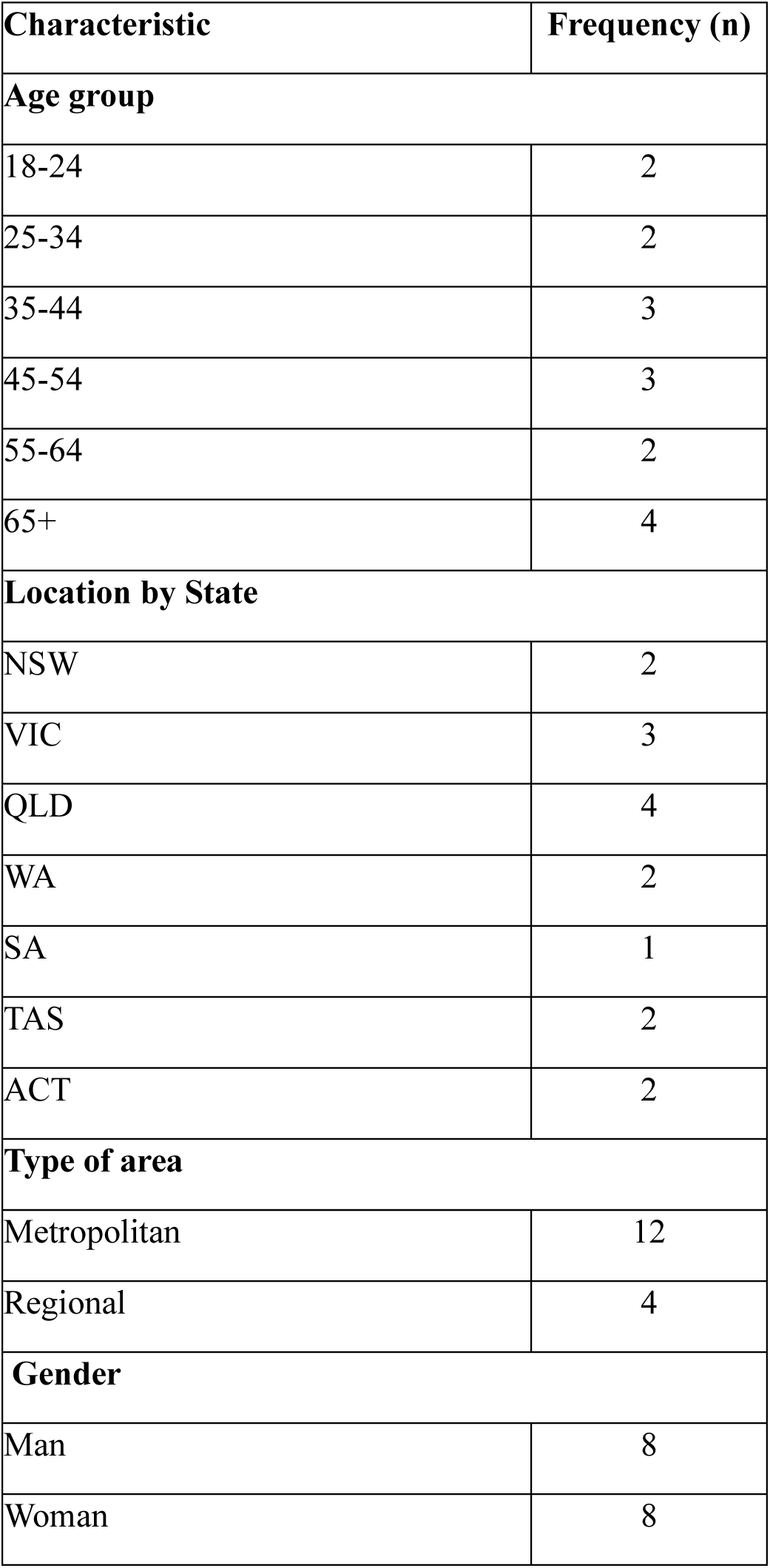
Summary of participant characteristics (n=16)

#### 3.1.1. Changes made to the survey items in Australia

We made changes to nine survey items based on cognitive interviews [see Additional file 1]. One survey item (SP5-B: what types of communication would you trust to give you accurate information about the influenza vaccine?) was removed as it did not measure trust in communication as intended. Five items were rephrased for various reasons including being not applicable to the Australian context (D7: education background), allowing answers from both those who received and those who had never received an influenza vaccine (VS2: place of receiving the vaccine), ambiguity in interpretations (VS3: payment made), inconsistencies with previous questions (SP4: recommendation from health worker), and unclear temporal references (M1: intention to get the vaccine).

We also added response options to three questions. We added two response options (Not sure and Not applicable) to SP3B (religious leaders’ influence) to better accommodate the Australian context where having no religion is common. We also added one response option (Not sure) to SP3 (community leaders’ influence) and one response option (The vaccine is not available) to P6 (Factors that make it hard for you to get an influenza vaccine) to reflect recurring participant suggestions.

### 3.2. Psychometric evaluation

#### 3.2.1. Participant characteristics summary

Following the cognitive interviews, the modified survey questions were answered by 2,055 Australian adults. Most of the participants lived in metropolitan areas (79%), followed by those in regional areas (15.5%). Most of the participants were from New South Wales (31.9%), followed by Victoria (28.6%) and Queensland (19%). About 52.1% of the participants were female, 47.7% were male, and 0.2% identified as non-binary. Participants’ age ranged from 18 to 92 years (M = 51.4, SD = 16). About one-third (33.7%) held a bachelor’s degree, followed by those with a trade certificate/apprenticeship and high school or equivalent (23.7% and 22.8%, respectively). The frequency distribution of the survey items are summarised [see Additional file 2].

#### 3.2.2. Criterion Validity

##### 3.2.2.1. Bivariate associations

Polychoric correlations were used to assess the correlation of the survey items with the intention to receive influenza vaccine. This method was chosen because all items are either ordinal or binary. As shown in Table 2, the correlation between the items with the intention to receive the influenza vaccine ranged from near-zero (*r*= 0.03 for SP7; decision autonomy) to a moderate correlation (*r*= 0.52 for SP1; responsibility to protect others).

**Table 2:** Bivariate polychoric correlations with an intention to receive influenza vaccine.

| Code | Item | Polychoric Correlation |
| --- | --- | --- |
| SP1 | Do you think you have a responsibility to get the influenza vaccine in order to protect others? | 0.52 |
| TF6 | How important do you think the influenza vaccine is for your health? | 0.44 |
| SP3 | Do you think most of your close family and friends want you to get the influenza vaccine? | 0.43 |
| **TF4** | How much protection do you think the influenza vaccine will offer you? | 0.42 |
| **TF5** | When a person gets the influenza vaccine, how much do you think it protects people in the community? | 0.42 |
| **TF7** | How safe do you think the influenza vaccine is for you? | 0.38 |
| **SP2** | Do you think most adults you know will get the influenza vaccine if it is recommended for them? | 0.36 |
| **TF3** | Before today, had you heard of the influenza vaccine? | 0.34 |
| **TF8** | How much do you trust the health workers who would give you the influenza vaccine? | 0.34 |
| **P03** | Do you know where to go to get an influenza vaccine for yourself? | 0.30 |
| **SP4** | Do you think your community leaders want you to get the influenza vaccine? | 0.28 |
| **TF1** | How concerned are you about getting seasonal influenza, also known as “the flu”? | 0.27 |
| **SP5** | In the last two years, has a health worker recommended that you get the influenza vaccine? | 0.24 |
| **SP4b** | Do you think your religious leaders want you to get the influenza vaccine? | 0.23 |
| **TF2** | How sick would you become if you had influenza? | 0.21 |
| **P06** | How easy is it for you to pay for the influenza vaccination? | 0.16 |
| **P07** | How easy is it to afford other costs associated with influenza vaccination? | 0.16 |
| **P01** | In the last 12 months, have you ever been contacted about being due for the influenza vaccine? | 0.13 |
| **P05** | How easy is it to get the influenza vaccine for yourself? | 0.13 |
| **SP7** | When it is time for you to get the influenza vaccine, who is the main decision maker? | - 0.03 |

##### 3.2.2.2. Multivariate Associations and Variable Importance

A random forest model was performed to identify the importance of each item in predicting the intention to receive influenza vaccine within the full set of items.

The random forest analysis (Figure 2) indicated that TF6 (vaccine importance) was most highly correlated with the intention to receive the influenza vaccine, with a polychoric correlation of 0.44. SP1 (responsibility to protect others) was second most highly correlated, although it had the highest polychoric correlation (*r*=0.52). SP4 (*r*= 0.28) and P03 (*r*= 0.30) taking third and fourth positions, respectively.

**Figure 2:**
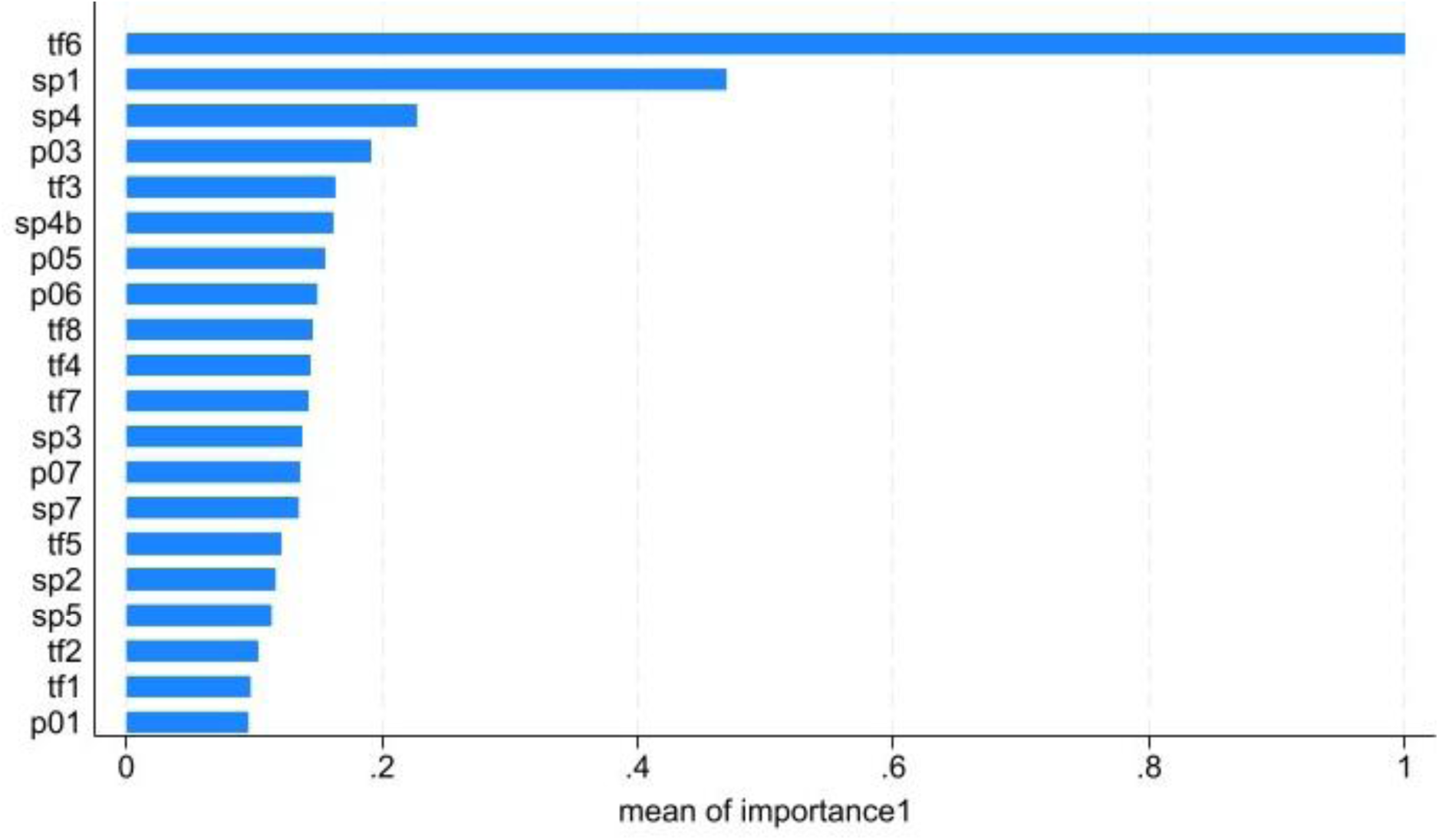
Importance of items in predicting intention to receive the influenza vaccine.

Although SP3 (family and friends influence; (*r*=0.43), TF4, and TF3 (awareness; *r*=0.42 for each) had moderately high correlations with the intention to receive the vaccine, the analysis did not identify them as the most important predictors. However, when considered together with the full set of items, they were found to be less important in predicting the intention to receive the influenza vaccine.

#### 3.2.3. Structure and model fit of the survey items

##### 3.2.3.1. Exploratory Factor Analysis

Exploratory Factor Analysis (EFA) was used to identify underlying constructs and understand how items were loaded on them. Five interpretable factors (i.e. constructs) with Eigenvalues greater than one were identified with oblique (Promax) rotation. Results from the 5-factor solution are shown in Table 3. Factor loadings mean how strongly an item is correlated with a particular construct.

**Table 3:** Factor loadings for 5-factor EFA solution.

| Code | Item | Beliefs | Access | Connection to community & resources | Perceived threat | Prompts |
| --- | --- | --- | --- | --- | --- | --- |
| TF1 | How concerned are you about getting seasonal influenza, also known as “the flu”? |  |  |  | .778 |  |
| TF2 | How sick would you become if you had influenza? |  |  |  | .869 |  |
| TF3 | Before today, had you heard of the influenza vaccine? |  |  | .593 |  |  |
| TF4 | How much protection do you think the influenza vaccine will offer you? | .805 |  |  |  |  |
| TF5 | When a person gets the influenza vaccine, how much do you think it protects people in the community? | .828 |  |  |  |  |
| TF6 | How important do you think the influenza vaccine is for your health? | .819 |  |  |  |  |
| TF7 | How safe do you think the influenza vaccine is for you? | .750 |  |  |  |  |
| TF8 | How much do you trust the health workers who would give you the influenza vaccine? | .587 |  |  |  |  |
| M1 | Do you want to get an influenza vaccine in the upcoming flu season (May-September 2024)? | .647 |  |  |  |  |
| SP1 | Do you think you have a responsibility to get the influenza vaccine in order to protect others? | .807 |  |  |  |  |
| SP2 | Do you think most adults you know will get the influenza vaccine if it is recommended for them? | .661 |  |  |  |  |
| SP3 | Do you think most of your close family and friends want you to get the influenza vaccine? | .780 |  |  |  |  |
| SP4 | Do you think your community leaders want you to get the influenza vaccine? |  |  | .821 |  |  |
| SP4b | Do you think your religious leaders want you to get the influenza vaccine? |  |  | .683 |  |  |
| SP5 | In the last two years, has a health worker recommended that you get the influenza vaccine? |  |  |  |  | .542 |
| SP7 | When it is time for you to get the influenza vaccine, who is the main decision maker? |  |  |  |  | .562 |
| P01 | In the last 12 months, have you ever been contacted about being due for the influenza vaccine? |  |  |  |  | .745 |
| P03 | Do you know where to go to get an influenza vaccine for yourself? |  |  | .407 |  |  |
| P05 | How easy is it to get the influenza vaccine for yourself? |  | .768 |  |  |  |
| P06 | How easy is it for you to pay for the influenza vaccination? |  | .910 |  |  |  |
| P07 | How easy is it to afford other costs associated with influenza vaccination? |  | .921 |  |  |  |

The first factor (Beliefs) was associated with nine items, including five (TF4-TF8), which were already categorised as *Thoughts and Feelings*; the *wanting a flu vaccine* (M1) item was considered to be a *Motivation* item, and three (SP1-SP3) were proposed to be *Social Processes* items. The second factor, feasibility and accessibility of getting influenza vaccine (Access), was associated with three items (P05-P07), which were considered to be *Practical Issues*. The third factor (the connection to community and resources) was correlated with practical knowledge of the influenza vaccine (TF3) and locations for getting the vaccine (P03), in addition to the influence of community and religious leaders (SP4 and SP4a). Two items (TF1 and TF2) were loaded on the fourth factor (Perceived threat) of the disease. The fifth factor (Prompts) loaded three items, including two items related to receiving recommendations and reminders from the healthcare system (SP5 and P0, respectively), in addition to one item (SP7), related to the respondent’s autonomy to make a decision about vaccination.

##### 3.2.3.2. Confirmatory Factor Analysis (CFA)

CFA was conducted by imposing restrictions on factor loadings to ensure model fit, indicating that most items were associated with only one underlying factor. CFA for a 4-factor model with the hypothesised structure (i.e., the BeSD framework with thinking and feeling, social processes, motivation and practical issues) was not performed because this model did not converge in the validation of the original BeSD [12]. We performed CFA to test the 5-factor model suggested by the EFA.

Table 4 shows factor inter-correlations. The strongest correlations were between Factor 1, Factor 5, and Factor 2 (r=0.518 and r=0.514), followed by the correlation between Factor 3, Factor 1 (r=0.498), and Factor 2 (r=0.466).

**Table 4:** Factor Inter-correlations.

|  | **Beliefs** | **Access** | **Connection to**<br>**community**<br>**and resources** | **Perceived**<br>**threat** | **Prompts** |
| --- | --- | --- | --- | --- | --- |
| **Factor 1** | 1 |  |  |  |  |
| **Factor 2** | 0.514 | 1 |  |  |  |
| **Factor 3** | 0.498 | 0.466 | 1 |  |  |
| **Factor 4** | 0.501 | 0.102 | 0.217 | 1 |  |
| **Factor 5** | 0.518 | 0.251 | 0.185 | 0.425 | 1 |

CFA showed the best goodness of fit for the survey with five factors (χ2=2939.3, p<0.001, RMSEA = 0.087, CFI = 0.855, TLI = 0.830). Table 5 shows all factor loadings were significant at p < .001 except SP7 (the main decision maker for the influenza vaccine) did not load significantly on Factor 5 (prompts) (*r*=0.032).

**Table 5:** Confirmatory Factor Analysis Results (Standardized)

|  | **Loading** |
| --- | --- |
| **Beliefs** |  |
| TF4 | .875 |
| TF5 | .854 |
| TF6 | .868 |
| TF7 | .799 |
| TF8 | .741 |
| M1 | .457 |
| SP1 | .714 |
| SP2 | .509 |
| SP3 | .667 |
| **Access** |  |
| P05 | .667 |
| P06 | .880 |
| P07 | .879 |
| **Connection to community**<br>**and resources** |  |
| TF3 | .512 |
| SP4 | .485 |
| SP4B | .554 |
| P03 | .602 |
| **Perceived threat** |  |
| TF1 | .735 |
| TF2 | .588 |
| **Prompts** |  |
| SP5 | .744 |
| SP7 | .032* |
| P01 | .386 |

## 4. Discussion

The cognitive interviews and the psychometric evaluation of the Behavioural and Social Drivers of the Influenza Vaccination survey in Australia confirmed its good validity.

Findings from cognitive interviews helped improve the validity of the survey items within the Australian context by enhancing their clarity and relevance. This highlights the need to adapt survey questions for specific cultural, social, and health system contexts before conducting the survey on a larger sample in order to enhance the generalizability of the survey’s findings. For instance, we added “Not sure” or “Not applicable” to the question about religious leaders. In responding to this question, participants often found a forced “yes” or “no” choice challenging since about 39% of Australians have no religion [13]. While adding a “no opinion” option should be avoided since it usually does not affect response quality and may lead to response bias in some population groups [14] and is difficult to treat in the analysis, it may be needed at times.

From the psychometric evaluation, we identified and validated five factors that underlie the survey structure. These include individual and social *beliefs* about the vaccine, *access* to the vaccine, *connection to community and resources* about the vaccine, *perceived threat* from the disease, and *prompts* to get the vaccine. They align well with the domains of the BeSD framework, Thinking and Feeling, Social Processes, Motivation and Practical Issues.

The findings also demonstrated each item correlated with the intention to receive influenza vaccine, particularly those related to perceived vaccine importance and social responsibility. Furthermore, the findings revealed the importance of each item in predicting the intention to receive influenza vaccine within the full set of items.

The social responsibility item was newly developed for this survey. WHO added it to measure the sense of responsibility to vaccinate as a pro-social motive [10]. A previous study explained the role of “collective responsibility,” which refers to an individual’s willingness to protect others, in vaccination behaviour [15]. Interestingly, this item performed well during the cognitive interviews and in the psychometric analysis, supporting its value as an addition to the survey tool.

Only one item, the SP7 (autonomy of decision-making about influenza vaccine), did not show a significant association with the intention to receive influenza vaccination (criterion validity), and also was not significantly correlated with its underlying factor (prompts) in the CFA. This finding was similar in the BeSD for childhood vaccination testing [12]. It therefore does not fit with the scale, but we recommend the retention of this question to provide descriptive information about the extent to which an adult is autonomous in decision-making, particularly in contexts where gender issues intersect with vaccination decision-making [16]. Furthermore, it may support the decisions about which groups should be targeted with recommendations in information campaigns. We also recommend validating the other versions of the survey in different countries.

## Limitation

We were not able to canvas all population groups in the Australian population. We surveyed very few First Nations people, and people who speak a language other than English would not have been able to take part in the cognitive interviews or survey. Also, remote communities and people experiencing other forms of social exclusion with limited access to the Internet were more likely to be under-represented. It was not possible to assess predictive validity because we did not measure subsequent 2024 influenza season vaccine uptake in this study. Validating against uptake would likely have produced different results since intention (wanting/motivation) is more likely to reflect perceptions and social influence than practical issues. Therefore, the findings of the study should be interpreted in light of the study’s context.

## Conclusion

The cognitive interviews and psychometric evaluation of the Behavioural and Social Drivers of the Influenza Vaccination survey showed good validity in an Australian population. These adaptations, like refining survey items for clarity and adding nuanced response options, were imperative in improving the generalizability of the findings. The survey’s underlying factors aligned well with the BeSD framework, particularly in measuring individual beliefs, social influences, and practical issues related to vaccination. Most items, including a newly developed social responsibility item, are significantly associated with the intention to receive the influenza vaccine, affirming their relevance in understanding vaccination behaviour. However, the item related to decision-making autonomy did not correlate significantly with its underlying factor nor with predicting vaccination intention, suggesting it may be more useful for descriptive purposes rather than as a predictive variable. An overall limitation of the validation was the inability to assess predictive validity due to the lack of prospective vaccination uptake data.

We recommend further validation of the survey in various cultural and health system contexts, with a particular focus on assessing its predictive validity by linking items to actual vaccination uptake following survey administration.

## Declarations

### Ethical approval and consent to participate

The University of Sydney Human Research Ethics Committee granted ethical approval. All research participants provided digital informed consent.

### Consent for publication

Not applicable.

### Availability of data and materials

The datasets used and/or analysed during the current study are available from the corresponding author upon reasonable request. Once the data are on the repository, we can refer any queries to the University of Sydney library dataset link.

### Authors’ contributions

MMS, MCE, and JL developed the survey protocol. MCE conducted and analysed the cognitive interviews. MCE led the survey’s data collection. MMS performed the psychometric analysis. MMS drafted the manuscript. JL reviewed and edited it. All authors have read and agreed to the published version of the manuscript.

## Acknowledgements

Not applicable.

